# A prospective cohort study of presenteeism and increased risk of unemployment among Japanese workers

**DOI:** 10.1101/2022.04.04.22273011

**Authors:** Yoshihisa Fujino, Makoto Okawara, Ayako Hino, Keiji Muramatsu, Tomohisa Nagata, Seiichiro Tateishi, Mayumi Tsuji, Akira Ogami, Tomohiro Ishimaru, the CORoNaWork project

## Abstract

**Objective:** We examined the association between presenteeism and risk of job resignations and unemployment among Japanese workers during the COVID-19 pandemic.

**Methods:** A prospective study of 27,036 Internet monitors was conducted, starting in December 2020, with 18,560 (68.7%) participating in the follow-up by December 2021. The Work Functioning Impairment Scale (WFun) was used to measure the degree of work function impairment.

**Results:** The group with the highest WFun scores had higher odds ratios (ORs) for both retirement and unemployment for health reasons than the group with the lowest WFun scores. ORs were 2.97 (95%CI: 2.46-3.59, p<0. 001) and 1.80 (95%CI: 1.64-1.98, p<0.001), respectively.

**Conclusions:** Workers with work functioning impairment were at increased risk of resignation or unemployment. Management strategies for workers with work functioning impairment are needed to reduce their disadvantages in employment.

## Introduction

Presenteeism is a condition in which a worker continues to work while in an unhealthy condition.[1] Two aspects of presenteeism are of public health concern. First, continuing to work while in poor health results in a loss of productivity, with consequential losses for the business.[2] In many regions, employers are generally at least partially responsible for the costs of health and medical care for their workers. They are therefore interested in the allocation of resources to invest in health support for workers, taking into account the direct costs of health and medical care as well as indirect costs such as reduced labor productivity and absenteeism.[3,4]

Second, presenteeism adversely affects workers’ quality of life, leading to further deterioration of their health and affecting their ability to continue working.[5,6] Working while in poor health means delays in treatment, missed opportunities for recuperation, exacerbation of symptoms, and increased risk of developing complications.[5–8] Eventually, the worker may find it difficult to continue in employment and be excluded from the labor market.[6,9] This has a significant impact on the economic situation of not only the workers themselves but also their families, leading to an increased burden on social security.

Viewed in the latter perspective, unemployment is one of the most serious consequences for workers experiencing presenteeism. A worker’s ability to work depends on the degree of mismatch between the health status, and job demands and work factors.[10,11] The extent to which an individual has difficulty working due to health reasons depends not only on the type and severity of disease, but also on a variety of factors, including the type of work, support of supervisors and co-workers, and the company’s health climate. A decrease in the worker’s ability to perform his or her job duties results in difficulties in remaining employed.[12,13]

The COVID-19 pandemic has affected workers’ experiences of presenteeism as well as their employment status. It has caused interruptions in treatment and increased anxiety due to infection among workers with chronic illnesses.[14–16] Lockdown and social distancing have resulted in public health challenges such as loneliness and stress.[17,18] These challenges lead to worker presenteeism. In addition, the COVID-19 pandemic is having a serious impact on the global economy and employment. Disasters often exacerbate the job insecurity of those whose health and socioeconomic status are vulnerable. We reported that during the COVID-19 pandemic, workers who were vulnerable in their socioeconomic status- - a high-risk group for unemployment--were more likely to experience presenteeism.[19] However, the association between presenteeism and unemployment among workers in the COVID-19 pandemic is not clear. The purpose of this study was to examine the association between presenteeism and resignations and unemployment during the COVID-19 pandemic.

## Materials and Methods

A prospective cohort study was conducted starting in December 2020 and followed up until December 2021. Participants completed questionnaires via the Internet at the beginning and end of the study. All participants gave informed consent, and the study was approved by the ethics committee of the XXXX (reference No. R2-079 and R3-006).

A protocol for this study was previously reported.[20] Participants were employed workers between the ages of 20 and 65 at the time of the baseline survey. Sampling was based on region, occupation, and sex. Regions were divided into five levels according to COVID-19 infection level, covering 47 prefectures. Occupations were divided into office workers and non-office workers. Finally, a total of 20 blocks were created, consisting of combinations of 5 regions, 2 occupations, and 2 sexes. Participants were included so that each block would have the same number of participants. The original plan was to collect data for 30,000 participants overall, with a minimum of 1,500 participants in each block.

The survey was commissioned to Cross Marketing Inc. (Tokyo, Japan). Approximately 600,000 of their 4.7 million pre-registered monitors were sent an email request to participate in the survey. Of these, 55,045 participated in the initial screening, with 33,302 satisfying the final inclusion criteria. Of those 33,302 participants, 27,036 were included in the analysis, after excluding those judged as submitting untrustworthy responses. The following criteria were used to determine untrustworthy responses: extremely short response time (≤6 minutes), reporting extremely low body weight (<30 kg), reporting extremely short height (<140 cm), inconsistent answers to similar questions throughout the survey (e.g., inconsistency on questions about marital status and area of residence), and wrong answers to a question used solely to identify unreliable responses (“Choose the third largest number from the following five numbers.”). Participants were required by the system to answer all questions, so there were no missing values. The follow-up survey was conducted in December 2021, one year after baseline. A total of 18,560 people (68.7%) participated in the follow-up after satisfying the inclusion criteria.

### Evaluation of work functioning impairment

The Work Functioning Impairment Scale (WFun) was used to measure the degree of work function impairment.[21] The WFun is a self-report outcome measure based on the Rash model, and its validity has been thoroughly verified according to consensus-based standards for the selection of health measurement instruments (COSMIN).[21–26] It consists of the following seven statements: “I haven’t been able to behave socially”, “I haven’t been able to maintain the quality of my work”, “I have had trouble thinking clearly”, “I have taken more rests during my work”, “I have felt that my work isn’t going well”, “I haven’t been able to make rational decisions”, and “I haven’t been proactive about my work.” The participant responds to each statement on a scale using a five-level scale: 1 = ‘not at all’, 2 = ‘one or more days a month’, 3 = ‘about one day a week’, 4 = ‘two or more days a week’ and 5 = ‘almost every day’. The WFun score thus ranges from 7 to 35 points: the higher the score, the greater the degree of work functioning impairment. Based on previous studies, a WFun score of 14 to 20 was considered moderately impaired and a score of 21 or higher was considered severely impaired work function.[26]

### Assessment of job loss, unemployment, and early retirement

At the follow-up survey, participants were asked about changes in their employment status: whether they had resigned and changed job due to health reasons, for other reasons not related to retirement, and whether they had been unemployed for any length of time during the one-year follow-up period.

### Other covariates

Information on participants’ characteristics was collected at baseline. Participants answered the following questions about themselves in an online form: age, sex, prefecture of residence, marital status (unmarried, bereaved/divorced), job type (mainly desk work, mainly involving interpersonal communication, mainly labor), number of employees in the workplace, educational background, income, smoking status, and alcohol consumption (6–7 days a week, 4–5 days a week, 2–3 days a week, less than 1 day a week, hardly ever).

### Statistical analyses

Age-sex adjusted odds ratios (ORs) and multivariate adjusted ORs were estimated using a multilevel logistic model nested in the prefecture of residence to take account of regional variability. We estimated ORs of work functioning impairment associated with resignation due to health reasons, due to other reasons unrelated to retirement, and experiencing unemployment, respectively. The multivariate model included age, sex, marital status, job type, income, education, smoking, alcohol drinking, number of employees in the workplace, and the incidence rate of COVID-19 by prefecture at baseline. A *p* value less than 0.05 was considered statistically significant. All analyses were conducted using Stata (Stata Statistical Software: Release 16; StataCorp LLC, TX, USA).

### Results

Compared to the group with the lowest WFun, the group with the highest WFun were of slightly higher average age (49.4 vs 46.6), were less likely to be married (58.6% vs 50.9%), and belonged to the lowest income groups (13.2% vs 19.1%). There were no reliable differences in occupational status or educational attainment, or in lifestyle habits such as smoking and drinking.

Table 2 shows the association of WFun with resignation experience and unemployment. WFun was associated with resignation due to health reasons. In the multivariate model, compared to the group with the lowest WFun, the OR of the group with a WFun of 14 points or more was 1.84 (96%CI; 1.48-2.28, p<0.001) and the OR of the group with a WFun of 21 points or more was 2.97 (95%CI: 2.46-3.59, p<0. 001).

**Table 1.**
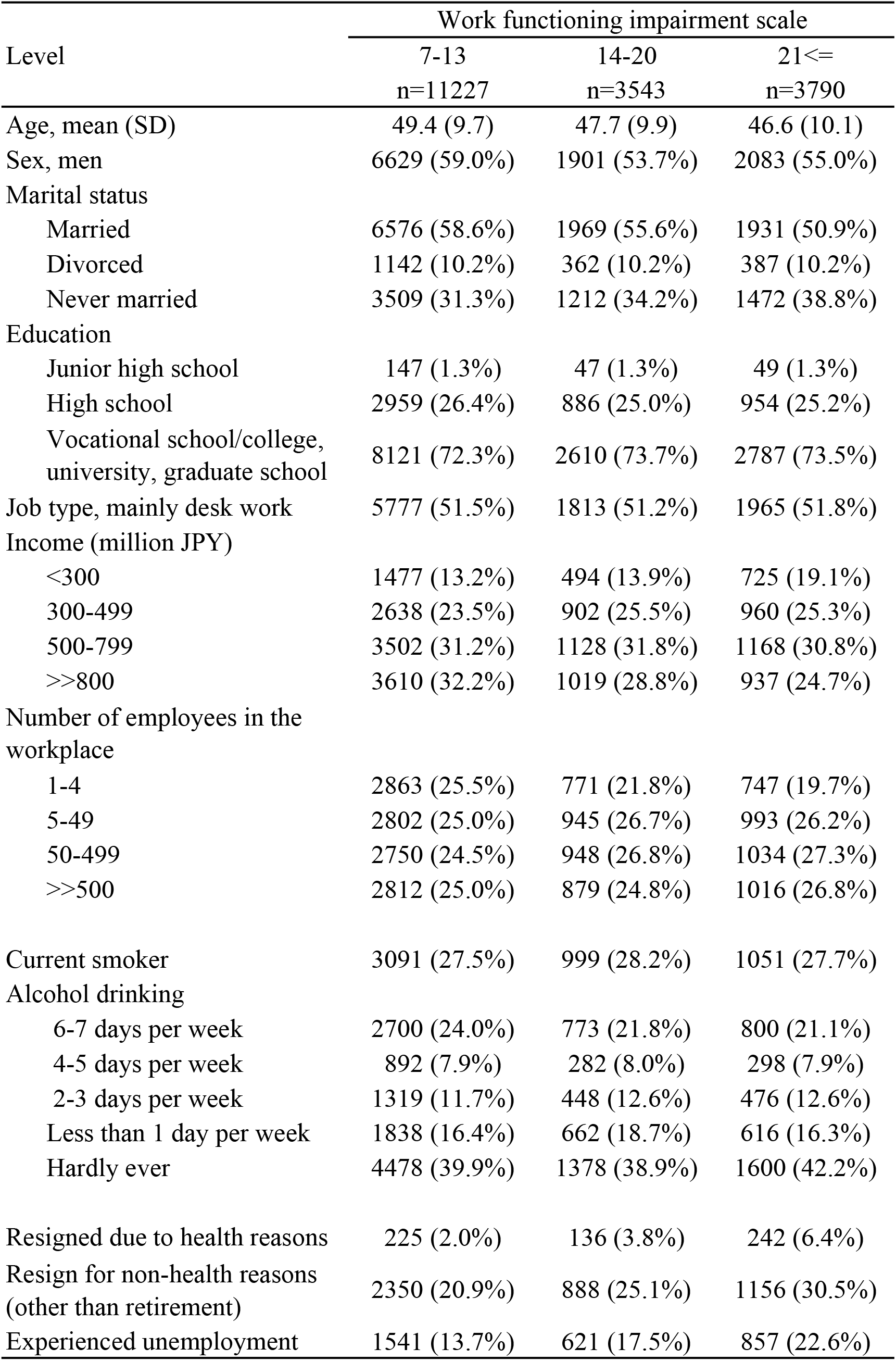
Baseline charactersitics according to WFun score level

| Level | Work functioning impairment scale |  |  |
| --- | --- | --- | --- |
|  | 7-13<br>n=11227 | 14-20<br>n=3543 | 21<=<br>n=3790 |
| Age, mean (SD) | 49.4 (9.7) | 47.7 (9.9) | 46.6 (10.1) |
| Sex, men | 6629 (59.0%) | 1901 (53.7%) | 2083 (55.0%) |
| Marital status |  |  |  |
| Married | 6576 (58.6%) | 1969 (55.6%) | 1931 (50.9%) |
| Divorced | 1142 (10.2%) | 362 (10.2%) | 387 (10.2%) |
| Never married | 3509 (31.3%) | 1212 (34.2%) | 1472 (38.8%) |
| Education |  |  |  |
| Junior high school | 147 (1.3%) | 47 (1.3%) | 49 (1.3%) |
| High school | 2959 (26.4%) | 886 (25.0%) | 954 (25.2%) |
| Vocational school/college,<br>university, graduate school | 8121 (72.3%) | 2610 (73.7%) | 2787 (73.5%) |
| Job type, mainly desk work | 5777 (51.5%) | 1813 (51.2%) | 1965 (51.8%) |
| Income (million JPY) |  |  |  |
| <300 | 1477 (13.2%) | 494 (13.9%) | 725 (19.1%) |
| 300-499 | 2638 (23.5%) | 902 (25.5%) | 960 (25.3%) |
| 500-799 | 3502 (31.2%) | 1128 (31.8%) | 1168 (30.8%) |
| >>800 | 3610 (32.2%) | 1019 (28.8%) | 937 (24.7%) |
| Number of employees in the<br>workplace |  |  |  |
| 1-4 | 2863 (25.5%) | 771 (21.8%) | 747 (19.7%) |
| 5-49 | 2802 (25.0%) | 945 (26.7%) | 993 (26.2%) |
| 50-499 | 2750 (24.5%) | 948 (26.8%) | 1034 (27.3%) |
| >>500 | 2812 (25.0%) | 879 (24.8%) | 1016 (26.8%) |
| Current smoker | 3091 (27.5%) | 999 (28.2%) | 1051 (27.7%) |
| Alcohol drinking |  |  |  |
| 6-7 days per week | 2700 (24.0%) | 773 (21.8%) | 800 (21.1%) |
| 4-5 days per week | 892 (7.9%) | 282 (8.0%) | 298 (7.9%) |
| 2-3 days per week | 1319 (11.7%) | 448 (12.6%) | 476 (12.6%) |
| Less than 1 day per week | 1838 (16.4%) | 662 (18.7%) | 616 (16.3%) |
| Hardly ever | 4478 (39.9%) | 1378 (38.9%) | 1600 (42.2%) |
| Resigned due to health reasons | 225 (2.0%) | 136 (3.8%) | 242 (6.4%) |
| Resign for non-health reasons<br>(other than retirement) | 2350 (20.9%) | 888 (25.1%) | 1156 (30.5%) |
| Experienced unemployment | 1541 (13.7%) | 621 (17.5%) | 857 (22.6%) |

**Table 2.** Odds ratios (ORs) of work functioning impairment associated with resignation due to health reasons, due to other reasons unrelated to retirement, and experiencing unemployment

| Outcome | WFun category | age-sex adjusted |  |  |  | multivariate* |  |  |  |
| --- | --- | --- | --- | --- | --- | --- | --- | --- | --- |
|  |  | OR | 95%CI |  | p | OR | 95%CI |  | p |
| Resigned due to health reasons | 7-13 | reference |  |  |  | reference |  |  |  |
|  | 14-20 | 1.87 | 1.50 | 2.32 | <0.001 | 1.84 | 1.48 | 2.28 | <0.001 |
|  | 21<= | 3.13 | 2.60 | 3.77 | <0.001 | 2.97 | 2.46 | 3.59 | <0.001 |
|  |  |  |  |  | <0.001<sup>†</sup> |  |  |  | <0.001<sup>†</sup> |
| Resigned for non-health reasons other than retirement | 7-13 | reference |  |  |  | reference |  |  |  |
|  | 14-20 | 1.25 | 1.14 | 1.36 | <0.001 | 1.24 | 1.13 | 1.36 | <0.001 |
|  | 21<= | 1.64 | 1.51 | 1.79 | <0.001 | 1.61 | 1.48 | 1.75 | <0.001 |
|  |  |  |  |  | <0.001<sup>†</sup> |  |  |  | <0.001<sup>†</sup> |
| Experienced unemployment | 7-13 | reference |  |  |  | reference |  |  |  |
|  | 14-20 | 1.35 | 1.22 | 1.49 | <0.001 | 1.33 | 1.20 | 1.47 | <0.001 |
|  | 21<= | 1.89 | 1.72 | 2.08 | <0.001 | 1.80 | 1.64 | 1.98 | <0.001 |
|  |  |  |  |  | <0.001<sup>†</sup> |  |  |  | <0.001<sup>†</sup> |

\* The model included age, sex, marital status, job type, income, education, smoking, alcohol drinking, number of employees in the workplace, and the incidence rate of COVID-19 by prefecture at baseline.
<sup>†</sup> p for trend

Similarly, WFun was also associated with resignation for other reasons unrelated to retirement. The multivariate model showed that the OR of the group with moderately impaired work functioning was 1.24 (95%CI: 1.13-1.36, p<0.001) and the OR of the group with severely impaired work functioning was 1.61 (95%CI: 1.48-1.75, p<0.001).

WFun was also associated with experience of unemployment. Comparing the group with moderate work functioning impairment and the group experiencing severe work functioning impairment, the ORs were 1.33 (95%CI: 1.20-1.47, p<0.001), and 1.80 (95%CI: 1.64-1.98, p<0.001), respectively.

## Discussion

The study showed that the higher the degree of work functioning impairment, the higher the risk of resigning from one’s job both due to health and non-health reasons. Furthermore, the higher the level of work functioning impairment, the higher the risk of experiencing unemployment. These results are consistent with previous reports that workers with illnesses such as rheumatoid arthritis, musculoskeletal disorders, mental health disorders, and cardiovascular disease, are associated with long-term sick leave and early retirement.[8,9,27,28]

Presenteeism is of growing interest to Japanese companies in accordance with the government-led so-called “health and productivity management strategy”.[29] However, this interest is generally focused on the decline in productivity caused by presenteeism,[10] a natural bias from the employer’s perspective given that reduced productivity is a strong business concern. In reality, however, presenteeism is not actively managed in Japanese companies in terms of long-term negative impacts on, for example, workers’ quality of life, and unemployment. Although in Japan routine health checkups are required by law, there is little incentive to actively manage presenteeism, which encompasses multiple health issues including symptoms and complaints that are sometimes not even diagnosed. In addition, workers who leave the workforce are unlikely to interest employers because the financial cost shifts from the company to somewhere else, e.g., social welfare.[30]

There are various possible reasons why resignations increase when workers experience presenteeism. People with health problems experience a variety of work disabilities.[13,31,32] Employees who are unable to perform certain tasks experience may have reduced work hours, changes in employment, and ultimately an increased risk of exiting the labor market.[13] Additionally, unhealthy individuals may have difficulty obtaining secure employment and are more likely to be employed by companies with unstable business conditions.[33,34] In such cases, they may be forced to resign or more likely to be made redundant independent of their health status.

While some illnesses are known to be associated with long-term sick leave and early retirement, [8,9,27,28] there is less evidence on the relationship between the degree of work functioning impairment and resignations in Japan.[35–38] Because the degree of work functioning impairment reflects a mismatch between job requirements and health status,[10,11] it cannot be evaluated solely on the basis of diagnosis or severity of a given disease. For example, a worker with back pain may be unable to tolerate muscle work but could cope with a desk job. In addition, evaluations based on, for instance, the glycosylated hemoglobin (HbA1c) for diabetes or the simplified disease activity index (SDAI) for rheumatoid arthritis, are appropriate for assessing disease severity, but they do not indicate the degree of difficulty a worker experiences while performing his or her job. Therefore, we argue for using multiple tools to measure presenteeism, for a comprehensive assessment of health-induced work difficulties in people with various illnesses and severities.

This study was conducted in Japan when the COVID-19 pandemic was expanding.[20] Many Japanese companies are implementing various infection control measures to prevent the spread of COVID-19, including maintaining physical distance, wearing masks, basic hygiene (e.g., hand washing), daily health checks, and telework. One measure was restricting employees from coming to work when they were sick. However, workers with concerns about their employment security may hesitate to report sickness to the company.[39–42] In addition, across the world interruptions to medical treatments have been caused by the COVID-19, including in Japan.[14] Interruptions and delays in treatment may exacerbate pre-existing health conditions and increase complications, which may further increase the incidence of sickness presenteeism. In parallel, the COVID-19 pandemic has led to precarious employment conditions in Japan.[43] During the pandemic, workers in a more insecure socioeconomic situation were more likely to experience presenteeism.[19] Vulnerable socioeconomic conditions are themselves also a risk factor for job resignation and unemployment. In this study, even after controlling for socioeconomic factors such as income, education, and firm size, work functioning impairments were still found to increase the risk of resigning and unemployment, thus creating a double whammy for those with socioeconomic insecurity.

Reducing employment disadvantage through early intervention in cases of work functioning impairment is a major challenge.[30,34] However, no specific scheme for this purpose has been forthcoming. Adjustment of the working environment is essential for people with health problems to continue working.[12] Workers’ health status and their ability to work can be modified to improve and maintain continuity of employment, for example, by adjusting workload and content, and by providing benefits such as sick leave for medical treatment and recuperation.[40,44,45] For some conditions, maintaining the ability to work has been achieved through clinical intervention.[46] However, many Japanese workers are known to avoid clinical intervention for pain and musculoskeletal disorders,[47,48] both conditions that can lead to presenteeism. Management of presenteeism requires both occupational and clinical support.

We note some limitations of this study. First, all the assessments were based on self-reports. However, we do not expect many serious memory errors or recall biases regarding changes in employment over the relatively short duration of the study (one year). Second, presenteeism and employment situations experienced by workers may be different under COVID-19 pandemic conditions than in a non-pandemic period. Concievably, the association could be even more pronounced. Third, we did not determine whether any period of unemployment was due to health reasons. Unemployment may occur simply because of the managerial situation of the employer. Nevertheless, because this study has revealed a greater likelihood of unemployment when the degree of work dysfunction is higher, the association may in fact be stronger than it appears.

In conclusion, workers with work functioning impairment for health reasons were at increased risk of resigning from their job, or unemployment. Obviously, the risk of resignation due to health reasons was particularly high. Further development of health management strategies for people with work functioning impairment is needed to reduce their disadvantages in regards to employment.

## Data Availability

All data produced in the present study are available upon reasonable request to the authors

## Acknowledgements

The current members of the CORoNaWork Project, in alphabetical order, are as follows: Dr. Akira Ogami, Dr. Ayako Hino, Dr. Hajime Ando, Dr. Hisashi Eguchi, Dr. Keiji Muramatsu, Dr. Koji Mori, Dr. Kosuke Mafune, Dr. Makoto Okawara, Dr. Mami Kuwamura, Dr. Mayumi Tsuji, Dr. Ryutaro Matsugaki, Dr. Seiichiro Tateishi, Dr. Shinya Matsuda, Dr. Tomohiro Ishimaru, and Dr. Tomohisa Nagata, Dr. Yoshihisa Fujino (present chairperson of the study group), and Dr. Yu Igarashi. All members are affiliated with the University of Occupational and Environmental Health, Japan.

